# The Genetic Landscape of Parkinson’s Disease in V4 countries of Central Europe - the CEGEMOD study

**DOI:** 10.64898/2026.06.30.26356950

**Authors:** Miriam Ostrozovicova, Alexandra Lackova, Milan Grofik, Petr Holly, Peter Klivenyi, Norbert Kovacs, Jan Necpal, Katarzyna Smilowska, Igor Straka, Gertrud Tamas, Agsha Atputhavadivel, Janette Baloghova, Jana Deptova, Petr Dusek, Vladimir Han, Marek Hornak, Robert Jech, Katarina Kalinova, Lucia Klimcakova, Kristina Kulcsarova, Egon Kurca, Hamin Lee, Veronika Magocova, Maria Marekova, David Murphy, Jana Neupauerova, Stefan Orkuty, Jana Papikova, David Pinter, Miroslava Rabajdova, Evzen Ruzicka, Tereza Serranova, Krisztina Soos, Tatiana Svorenova, Peter Valkovic, Katerina Zarubova, Zuzana Gdovinova, Mie Rizig, Henry Houlden, Matej Skorvanek, the CEGEMOD consortium

## Abstract

**Background:** Parkinson’s disease (PD) genetics has been predominantly studied in Western European and North American populations, leaving Central Europe underrepresented. We aimed to characterise the genetic architecture of PD in the V4 countries (Slovakia, Czech Republic, Hungary, and Poland).

**Methods:** The CEGEMOD study combined a systematic review of published PD genetic studies from the V4 region with prospective genetic screening of 1373 patients. All participants underwent genotyping array screening, while genetically unresolved patients with early-onset or familial PD underwent whole-exome sequencing analysis. Variants were interpreted using current clinical standards and validated using Sanger sequencing.

**Results:** The systematic review identified 48 eligible studies reporting pathogenic or risk variants in PD patients from the V4 countries. In the prospective cohort, pathogenic, likely pathogenic, or established risk variants were identified in 157/1,373 patients (11.4%). *GBA1* variants accounted for the majority of findings, followed by *LRRK2, PRKN, POLG, PLA2G6*, and *ATP1A3*. Whole-exome sequencing provided an additional 5% diagnostic yield in genetically unresolved high-risk patients. Our findings also demonstrate substantial disparities in genetic research across Central Europe.

**Conclusions:** This study provides the largest genetic characterisation of PD in Central Europe to date. The CEGEMOD study expands knowledge of the regional genetic landscape, supports the implementation of genetic testing in clinical practice, and establishes an important resource for future precision medicine and collaborative PD genetics research.

## Introduction

Parkinson’s disease (PD) is the second most common neurodegenerative disorder, with a complex genetic aetiology, which has also been acknowledged by the newly defined biologically based classification of PD (Höglinger et al. 2024). The genetic variants are estimated to contribute approximately 25% to the overall risk of developing PD; however, they vary in terms of frequency and disease risk. Approximately 5-10% of individuals with PD carry rare variants in single genes that can cause disease (i.e. are pathogenic) following the classical Mendelian inheritance pattern, predominantly discovered through linkage analysis of affected families (Day and Mullin 2021). Understanding of the genetic mechanisms underlying the disease is a crucial step towards the development of targeted treatments. Therefore, the improved availability of genetic testing and precision medicine leads to an urgent need for expert consensus on determining the causal genes and variants for PD.

However, despite these advances, the global PD genetic data generated so far are currently limited and focus mostly on European and North American populations (Schumacher-Schuh et al. 2022; Towns et al. 2023). This significant lack of population diversity in PD research highlights the immediate need for better representation. The ‘‘underrepresented’’ populations are now being recognised, with international groups that aim to accelerate the identification of novel genetic causes of PD, e.g. within the partnership of Global Parkinson’s Genetics Programme (GP2) (Lange et al. 2023).

Lack of ethnic diversity is a known systematic issue in genetic studies in general, and even though majority of the genomic data in research are coming from Europe, these mostly represent populations of developed countries of the northwestern regions, and the uneven distribution of the available data is further impaired by the lack of data sharing, and the lack of comprehensive computational and storage infrastructure to facilitate data processing in clinically relevant timeframe (Bagger et al. 2024). While we tend to think of Europeans as one single group, genetically, there has already been shown a high degree of variability. Recent whole-genome studies have already demonstrated that local genome diversity and population structure in Eastern European regions have not been fully represented, underscoring the importance of closing the remaining gaps in human genome diversity (Oleksyk et al., 2022).

The aim of this study is, therefore, in collaboration with the CEGEMOD consortium, to characterise the genetic architecture of PD in countries of Central Europe, identify and describe the frequency of pathogenic Mendelian gene variants, novel rare variants or risk variants for PD and to describe the overlap in the genetic architecture of PD in Central Europe compared to other European and Asian ancestries. Overall, we wish to increase the diversity in PD genetics data to improve our understanding of genetic architecture in all populations, with the potential to target genetic testing and drug development.

## Material and methods

### Systematic Review

The systematic review of genetic studies conducted so far in the V4 countries of Central Europe followed the PICOS (population, intervention, comparison, outcome, and study type) framework (Amir-Behghadami and Janati, 2020) (Supp. Table 1) and Preferred Reporting Items for Systematic Reviews and Meta-analyses (PRISMA) guidelines (Page et al., 2021). In brief, the literature was comprehensively searched and identified in the following English databases: MEDLINE/PubMed, Web of Science and Scopus. The full search syntax without any date or language limitations is presented in Supp. Table 2. Results were first screened based on a rapid review of titles and abstracts, and irrelevant studies were excluded. Potentially relevant studies were obtained in full and reviewed in detail. This review included all observational studies, single case reports, and case series investigating genetic aspects of PD in patients diagnosed by either the UK Parkinson Disease Society (UKPDS) brain bank or the Movement Disorders Society (MDS) diagnostic criteria, without any date limitations. Studies in either English or the native language of each country were included. Studies written in the native language were reviewed, included and rated by the native speaker specialists. Studies involving only genes not established as causes of monogenic PD (based on the Genomics England Parkinson Disease and Complex Parkinsonism panel list v1.120 - (https://panelapp.genomicsengland.co.uk/panels/39), MDSGene online database (https://www.mdsgene.org/) (Lill et al., 2016) and published literature (Blauwendrat et al, 2020, Gustavsson et al., 2024, Magrinelli et al., 2026) ), non-human studies and in vitro studies were excluded. Participants with origins outside the V4 countries of Central Europe (Czech, Poland, Slovakia and Hungary) were excluded. An overview of the literature search as well as the filtering procedure is displayed in Supp. Fig. 1, and the list of all eligible articles can be found in Supp. Table 3. The following data were extracted: candidate genes and single-nucleotide polymorphisms (SNPs) or copy number variations (CNVs), identified candidate genes/SNPs/CNVs, investigated study design, population characteristics including ethnicity, sample sizes in each study group, genotyping method, clinical interpretation, lead author, country and year of publication. We used the p. amino acid change identifier using the dbSNP database (https://www.ncbi.nlm.nih.gov/snp). When unsuccessful, we have reported the variant name as it was reported by the original study authors. To ensure up-to-date assessment of variant pathogenicity, all identified variants were additionally re-evaluated using the ClinVar database (https://www.ncbi.nlm.nih.gov/clinvar/). In order to make the information openly accessible, we built the Parkinson Disease Variant Catalogue of V4 countries (https://miriam-ostrozovicova.shinyapps.io/Systematic_Review_PDrare/). This browser was built using R Shiny and includes specific information about each variant reported as follows: lead author and year of the manuscript that reported the variant, manuscript’s PMID, gene name, variant name, number of identified PD carriers, number of total PD patients screened, zygosity of the identified PD carriers, number of identified HC carriers, number of total HC screened, zygosity of the identified HC carriers, ethnicity of those carriers, clinical interpretation of the variant stated in the manuscript, up-to-date re-evaluation of the clinical interpretation using ClinVar, and inheritance of identifed variant in PD based on OMIM. The data were generated and compiled before being added to the browser.

### Participants

The CEGEMOD (Central European group on GEnetics of MOvement Disorders) study is a multi-centre clinical study conducted in four Central European countries (historically known as the Visegrad Group, or V4): the Slovak Republic, the Czech Republic, Poland, and Hungary. All participants were recruited voluntarily and gave written informed consent before their enrolment. This study was approved by the ethics committees of all involved institutions. Each individual with PD was diagnosed in accordance with the MDS clinical diagnostic criteria (Postuma et al. 2015). After the enrolment, detailed demographic and clinical data, as well as peripheral venous blood for DNA extraction, were collected using a standardised protocol as described elsewhere (Ostrozovicova et al., 2024; Ostrozovicova et al., 2025).

### Sample collection and DNA extraction

Peripheral venous blood was used to collect whole blood samples, which were then stored at -20°C or -80°C based on local capacities. Whole blood samples were defrosted on ice and then used for genomic DNA (gDNA) isolation using the QIAamp DNA Blood Mini kit (Qiagen, Hilden, Germany) according to the valid methodological protocol approved by the kit manufacturer. The concentration and quality control of purity in the isolated gDNA samples were tested using Nanodrop LC 3000 (ThermoScientific, Bratislava, Slovak Republic) in accordance with the manufacturer’s instructions and gDNA concentration was adjusted to appropriate values depending on the following genetic analyses as described elsewhere (Ostrozovicova et al., 2025).

### Genetic testing and the Definition of gene set

The genetic study has had 2 phases: a pilot study (Phase 1), where all recruited participants were first screened for known pathogenic, likely pathogenic or risk variants within the PD-related genes using GSA-24 v.3.0 Illumina array. In the second phase (Phase 2), whole-exome sequencing (WES) was used in a subgroup of participants diagnosed with PD based on MDS-PD diagnostic criteria and previous negative genetic finding on GSA-24 v3.0 microarray based on the following inclusive criteria: 1. early-onset defined as an age of onset (AOO) below 50 (1 point if AOO < 50; 2 points if AOO < 40); 2. AND/OR positive family history (1 point; defined as having at least one other affected relative in the family); 3. AND/OR presence of consanguinity (2 points). The scoring system was applied, with participants reaching a minimum of 2 points being prioritised for the WES analysis.

The definition of gene sets was based on Genomics England Parkinson Disease and Complex Parkinsonism panel list v1.120 - green entities (https://panelapp.genomicsengland.co.uk/panels/39) and published literature (Blauwendrat et al, 2020; Gustavsson et al., 2024; Magrinelli et al., 2026). We included both validated genes causing PD or atypical parkinsonism’ syndromes with Mendelian inheritance, as well as *LRRK2* and *GBA1*, as established risk factors. PD patients who tested negative in Phase 2 were subsequently included in an exploratory Phase 3 analysis. In this final phase, the virtual gene panel was expanded to include all genes classified as ‘amber’ or ‘red’ in the Genomics England Parkinson Disease and Complex Parkinsonism panel list v1.120 or curated as ‘green entities’ in the Genomics England Adult-onset neurodegenerative disorder panel list version 8.0. (https://nhsgms-panelapp.genomicsengland.co.uk/panels/474/v8.0), in order to assess not-yet validated PD-genes as well as potential phenotypic PD-mimics (Supp. Table 4).

### Genetic analysis - Phase 1

First, we performed SNPs genotyping using the GSA-24 v.3.0. to access the known pathogenic variant within the PD-related genes (Supp. Table 4) as described elsewhere (Ostrozovicova et al., 2025). The genotyping data were then generated in the raw IDAT files that were imported to GenomeStudio 2.0 software and underwent several quality-control procedures (QCs) to ensure the data quality, minimise systematic bias and prevent the occurrence of false positive or false negative results.

The sample QC steps were first used to identify contaminated samples. Samples with the genotyping call rate below 0.98 (98%) were excluded. The rate of heterozygosity was calculated across the entire dataset, and samples with excessive or deficient proportions of heterozygote genotypes, suggesting the possibility of gDNA sample contamination, were excluded (F cut-off of -0.15 and <-0.15 for inclusion). To identify the discrepancies between genotyped data and the recorded sex for each sample, the homozygosity rate across all X-chromosome SNPs was calculated and compared with the expected value. If the homozygosity rate (F value) was higher than 0.25 but less than 0.75, it suggested that the genotyping data had unclear sex information and that the sample was removed from downstream analysis unless it was confirmed that the gender data had been incorrectly recorded. Duplicated samples were removed using identity-by-descent calculation. The relatedness was estimated by calculating the pi-hat ratio, where values of 1 indicated duplicate samples, which were removed if not confirmed by the clinician as identical twins. On a variant QC level, a GenTrain Score of 0.80 was used as a cut-off for the measurement of SNP calling quality. GenomeStudio 2.0, PLINK version 1.9 and GCTA were used for the QC pipeline described above.

For variant selection, we excluded variants with a minor allele frequency (MAF) > 5%, non-coding variants and variants reported as benign or likely-benign on ClinVar (accessed April 2026). Using cleaned data, PLINK1.9 was used to filter out all the variants within the PD-associated genes’ in-house list (Supp. Table 4) and calculate the zygosity of positive SNPs. All positive SNPs were then also visually re-evaluated in the GenomeStudio 2.0 software.

### Genetic analysis - Phase 2

WES was performed by Novogene (Cambridge, UK) according to their protocol (Modi et al. 2021). The libraries were sequenced with Illumina’s NovaSeq 6000. All samples passed the internal QC steps with an average of 6.7 million total reads per sample and an average sequence depth of 33.7±4.2 x. FASTQ data were aligned against the GRCh38 human reference assembly using the IoN Queen Square in-house bioinformatic pipeline as described elsewhere (Ostrozovicova et al., 2025).

Following alignment, variants were filtered using specific thresholds for several annotations using the custom in-house pipeline. Synonymous and common variants with a population allele frequency (AF) ≥0.0001 (0.01%) reported in the Genome Aggregation Database v4.1 (gnomAD v4.1); Exome Aggregation Consortium (ExAC) database, or reported to be common in the IoN Queen Square in-house WES database, as well as variants predicted to be of functionality ‘low impact’ were removed. Candidate variants were grouped into four main functional categories. Loss-of-function variants included frameshift, stop-gained, canonical splice acceptor, canonical splice donor, start-lost, stop-lost, transcript-ablation, exon-loss, and protein-altering consequences. Missense variants were retained if they were rare and internally uncommon and had at least one supportive pathogenicity feature: SIFT annotated as deleterious, PolyPhen annotated as damaging, CADD PHRED score ≥20, or a ClinVar classification containing “pathogenic” or “likely pathogenic.” Splice-related variants were retained if the consequence annotation contained “splice” or if any SpliceAI delta score was greater than 0.5. In addition, ClinVar pathogenic or likely pathogenic variants were rescued under the relaxed rarity threshold. Lastly, only variants within the in-house gene panels were included for each phase, respectively (Supp. Table 4).

Final candidate variants were manually reviewed for gene-disease validity, inheritance pattern, zygosity, read depth, variant quality metrics, and consistency with the patient phenotype. In case of enough DNA material, the SNP variants which met these criteria and were considered as disease-causing were validated either by Sanger sequencing (for the SNPs and small CNVs), or by the targeted long-read sequencing (LRS) using the Oxford Nanopore MinION (for the heterozygous *GBA1* variants) (Ip et al. 2015). Variants of unknown significance (VUS) were not disclosed to participants, but were catalogued for future research use.

### Variant validation

Primers for mutation validation by Sanger sequencing were designed using Primer3 software (Rozen and Skaletsky 2000). Reference sequences of the human genome assembly build 38 (GRCh38) were obtained from the Ensemble Genome Browser (http://www.ensembl.org/index.html). The Primer-BLAST online tool (https://www.ncbi.nlm.nih.gov/tools/primer-blast/) was used to ensure the designed primers were specific to the target of interest, and the PCR reaction was then optimised and confirmed through agarose gel electrophoresis. The purified PCR product (Exosap, Thermo Fisher Scientific, USA) was then sequenced and read by the ABI 3730xl DNA analyser (Applied Biosystems, USA). The sequencing reads were checked using Sequencer software version 4.1.4.

The GBA1 variants were validated using a modified amplification protocol (Cuconato, G. et al., 2025), followed by long-read sequencing on the MinION Oxford Nanopore Technologies (ONT) platform when DNA of sufficient quantity and quality was available. The GBA1 region was enriched by amplification of a 6.5 kb long-range PCR product encompassing all coding exons, introns, and part of the 3′UTR. PCR amplification of the GBA1 target region was performed using optimised cycling conditions (94 °C for 2 min; 30 cycles of 94 °C for 30 s, 62 °C for 30 s, and 65 °C for 7 min 30 s; followed by 72 °C for 10 min) and 2 x Taq Master Mix (Vazyme Biotechnology Pty. Ltd., Canada). The primer sequences used were GBA1_6.5 kb_Forward: 5′-TCCTAAAGTTGTCACCCATACATG-3′ and GBA1_6.5 kb_Reverse: 5′-TAGTCACAGACAGCGTGTGAGC-3′. Amplicons were purified using VAHTS DNA Clean Beads (Vazyme Biotechnology Pty. Ltd., Canada), and DNA concentration was quantified using a Qubit 4 Fluorometer. For sample multiplexing, barcoding was performed using the Rapid Barcoding Kit (Oxford Nanopore Technologies, UK) according to the manufacturer’s amplicon sequencing protocol. Flow cell priming and sequencing were performed on a MinION R10.4.1 flow cell (FLO-MIN114). Sequencing was performed using the ONT platform with MinKNOW v25.09.16. Data acquisition and processing utilised Bream v8.8.3, Configuration v6.8.9, Dorado v7.11.2, and MinKNOW Core v6.8.11.

## Results

### Systematic Review

In total, the search resulted in 667 publications (Supp. Figure 1). After evaluating in a two-step procedure, a rapid screening of titles and abstracts, followed by full text, 48 publications contained information on PD mutation carriers from the V4 countries of Central Europe that were eligible for inclusion in the database. 19 from Poland (six as case reports), 8 from Hungary (two as case reports), 8 from the Czech Republic (two as case reports) and three from Slovakia. Furthermore, ten multicentric studies were conducted in collaboration with two or more countries (Supp. Table 5). Altogether, 74 *GBA1*, 34 *PRKN*, 22 *LRRK2*, 6 *PINK1*, 4 *PLA2G6*, 3 *DCTN1*, 1 *POLG*, 1 *SNCA*-associated PD patients were identified carrying already known pathogenic or likely pathogenic mutation in a heterozygous (for *GBA1, LRRK2, SNCA*) or homozygous/compound heterozygous (for *PRKN, PINK1, PLA2G6, POLG*) state (Figure 2), after excluding the monogenic PD carriers of unknown ethnicity reported in one multicentric study (Supp. Table 5), and are also available online at the Parkinson Disease Variant Catalogue of V4 countries (https://miriam-ostrozovicova.shinyapps.io/Systematic_Review_PDrare/).

**Figure 1.**
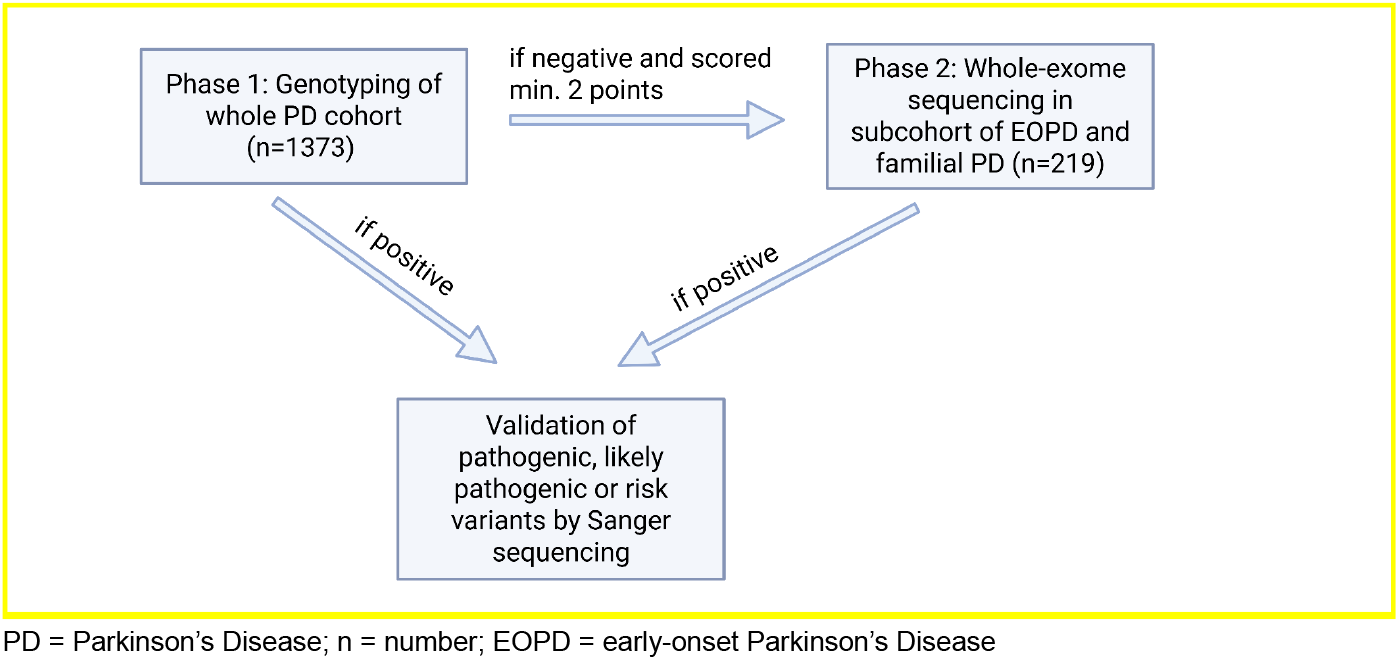
Study workflow

**Figure 2.**
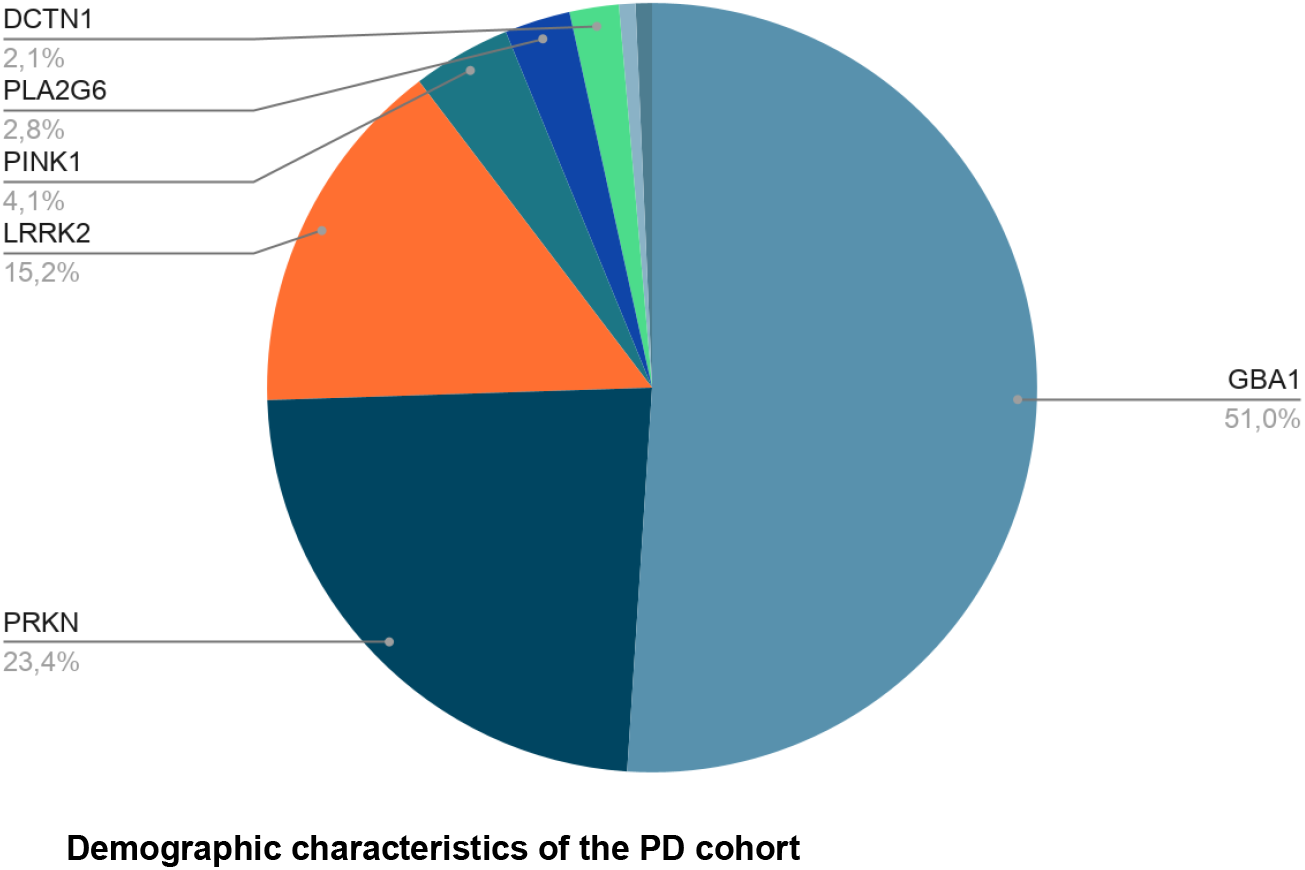
Graphical representation of published PD patients from V4 countries with pathogenic, likely pathogenic or risk variant in PD-associated gene

**Figure 2.**
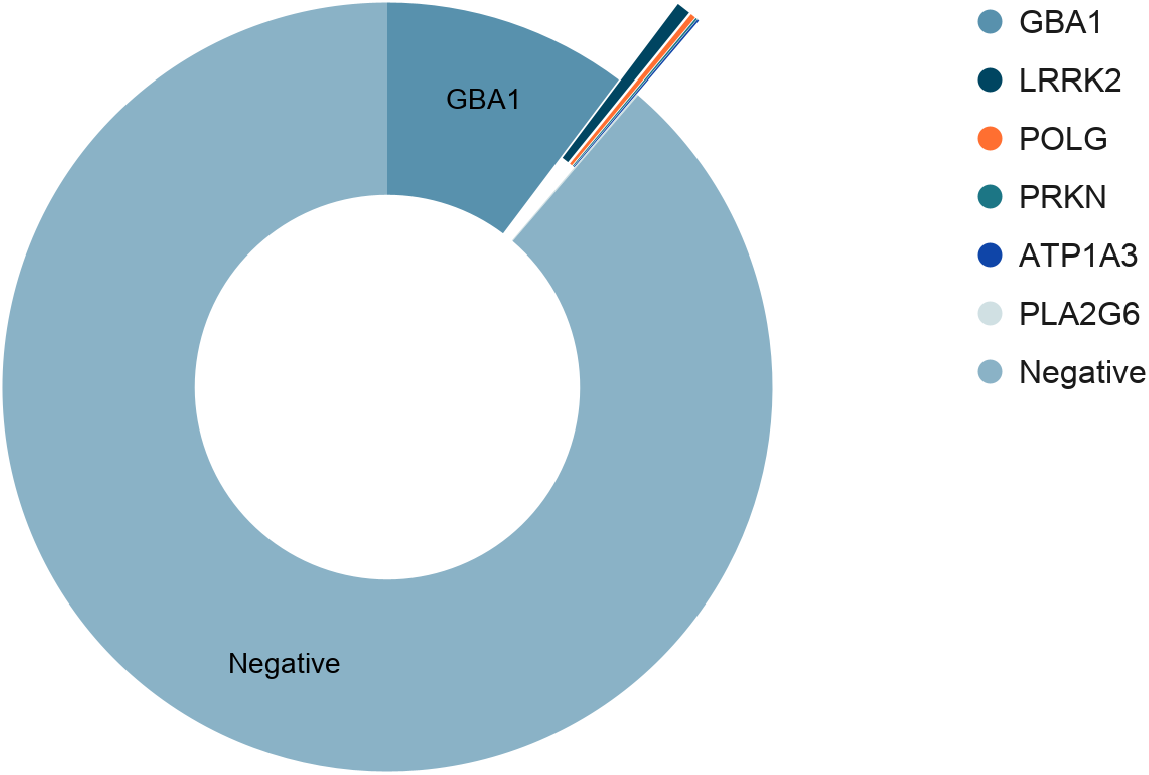
Variant level results

#### Demographic characteristics of the PD cohort

The mean age of PD patients included from Central Europe (n=1373), who had passed the quality control of the data, was 67.2±11.1 years, of whom 796 (58%) were men. Family history was positive in 195 patients (14.2 %), and 117 patients (8.5 %) developed PD before the age of 40 years. After first-line screening with genotyping data, 219 PD patients scored the required minimum of 2 points in the scoring system and were prioritised for second-line screening with WES. The mean age was 53.5±12.9 years, of whom 136 (61.8%) were men. Family history was positive in 93 patients (42.3%), and 117 patients (53.2%) developed PD before the age of 40 years. The basic characteristics of the patients included are summarised in Table 2.

**Table 2.** Demographic characteristics of the PD cohort.

| Study cohort | Genotyping array<br>(n=1373) | WES (n = 219) |
| --- | --- | --- |
| **Age** |  |  |
| **Mean±SD** | 67.2±11.1 | 53.5±12.9 |
| **Median** | 69 | 51 |
| **Gender (n, %)** |  |  |
| **Male** | 796 (58%) | 136 (62.1%) |
| **Female** | 577 (42%) | 83 (37.9%) |
| **Ethnicity (n, %)** |  |  |
| **Czech** | 207 (15.1%) | 56 (25.6%) |
| **Hungarian** | 245 (17.8%) | 43 (19.6%) |
| **Polish** | 31 (2.3%) | 19 (8.7%) |
| **Slovak** | 892 (65%) | 101 (46.1%) |
| **Age at onset (Mean±SD)** | 57.62±12.3 | 41.7±11.3 |
| **< 40 years (n, %)** | 117 (8.5%) | 117 (53.2%) |
| **< 50 years (n, %)** | 334 (24.3%) | 180 (81.8%) |
| **Positive family history (n, %)** | 195 (14.2%) | 93 (42.5%) |

PD = Parkinson's Disease; n = number; SD = standard deviation

#### Genetic findings

Reportable pathogenic, likely pathogenic or risk variants were identified in 157 participants. The overall yield for disease-relevant results, e.g. eliminating those who were heterozygous for autosomal recessive genes such as *PRKN, PINK1, DJ-1*, and adding those individuals who had common mild risk *GBA1* p.(Thr408Met) and p.(Glu365Lys) variants, was overall 11.4% (157 from 1373). Demographic and clinical data of all carriers are summarised in Table 3.

Using genotyping data within the Phase 1, we first identified *GBA1* risk variants in 139 patients (one patient was heterozygous for p.(Arg398Ter), five heterozygous for p.(Gly416Ser), 19 heterozygous carriers of the p.(Asn409Ser) variant, 34 were heterozygous for the p.(Glu365Lys), 77 were heterozygous and three homozygous for the p.(Thr408Met)). The second most common were *LRRK2* mutations in four patients (all as p.(Gly2019Ser) heterozygous carriers), *POLG* homozygous p.(Gly1051Arg) and *PLA2G6* homozygous p.(Ala633Val) mutation in one patient, respectively. Regarding the atypical Parkinsonism genes, one PD patient was positive for the pathogenic *ATP1A3* variant p.(Glu324Gly). Leveraging WES data of the previously genetically negative patients within the Phase 2 yielded additional *LRRK2* pathogenic mutation carriers (one heterozygous p.(Asn1437Ser) and 3 heterozygous p.(Leu1795Phe) carriers reported previously (Ostrozovicova et al. 2025), four patients were positive for *GBA1* risk variants (two for p.(Pro68ArgfsTer23), one heterozygous for the p.(Leu483Pro) and one for p.(Thr362Ile), two patients were double heterozygous carriers for *POLG* (both p.(Pro587Leu/p.Thr251Ile) variants) and one *PRKN* homozygous carrier p.(Arg42Pro). All findings are summarised in Figure 2 and Table 4.

**Table 3.**
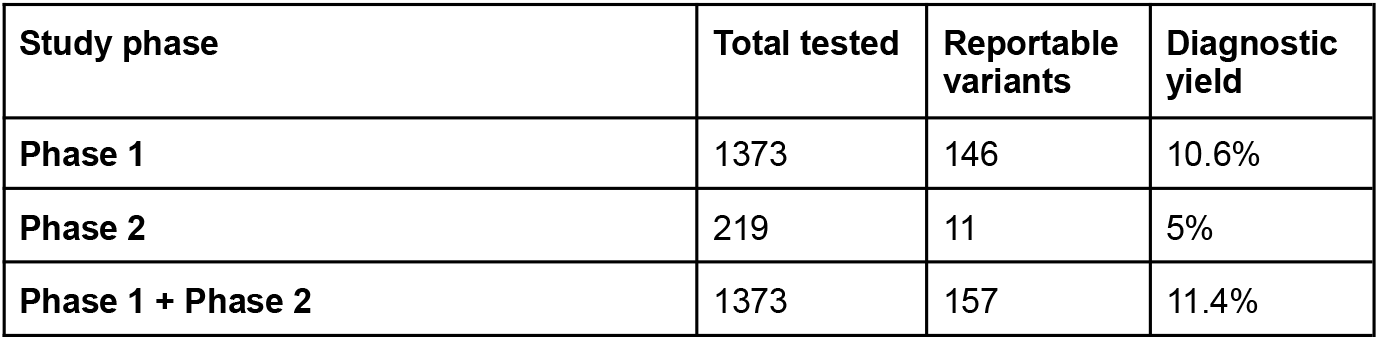
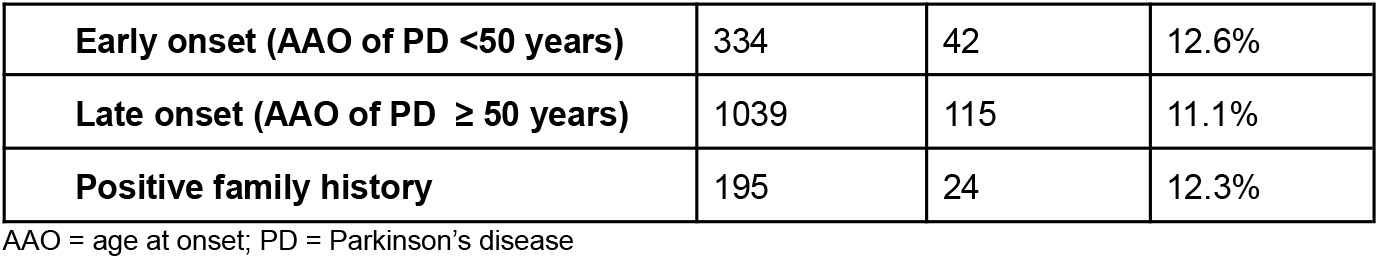
Genetic yield by study phase and by population.

| Study phase | Total tested | Reportable variants | Diagnostic yield |
| --- | --- | --- | --- |
| **Phase 1** | 1373 | 146 | 10.6% |
| **Phase 2** | 219 | 11 | 5% |
| **Phase 1 + Phase 2** | 1373 | 157 | 11.4% |
| **Early onset (AAO of PD <50 years)** | 334 | 42 | 12.6% |
| **Late onset (AAO of PD ≥ 50 years)** | 1039 | 115 | 11.1% |
| **Positive family history** | 195 | 24 | 12.3% |

AAO = age at onset; PD = Parkinson's disease

**Table 4.**
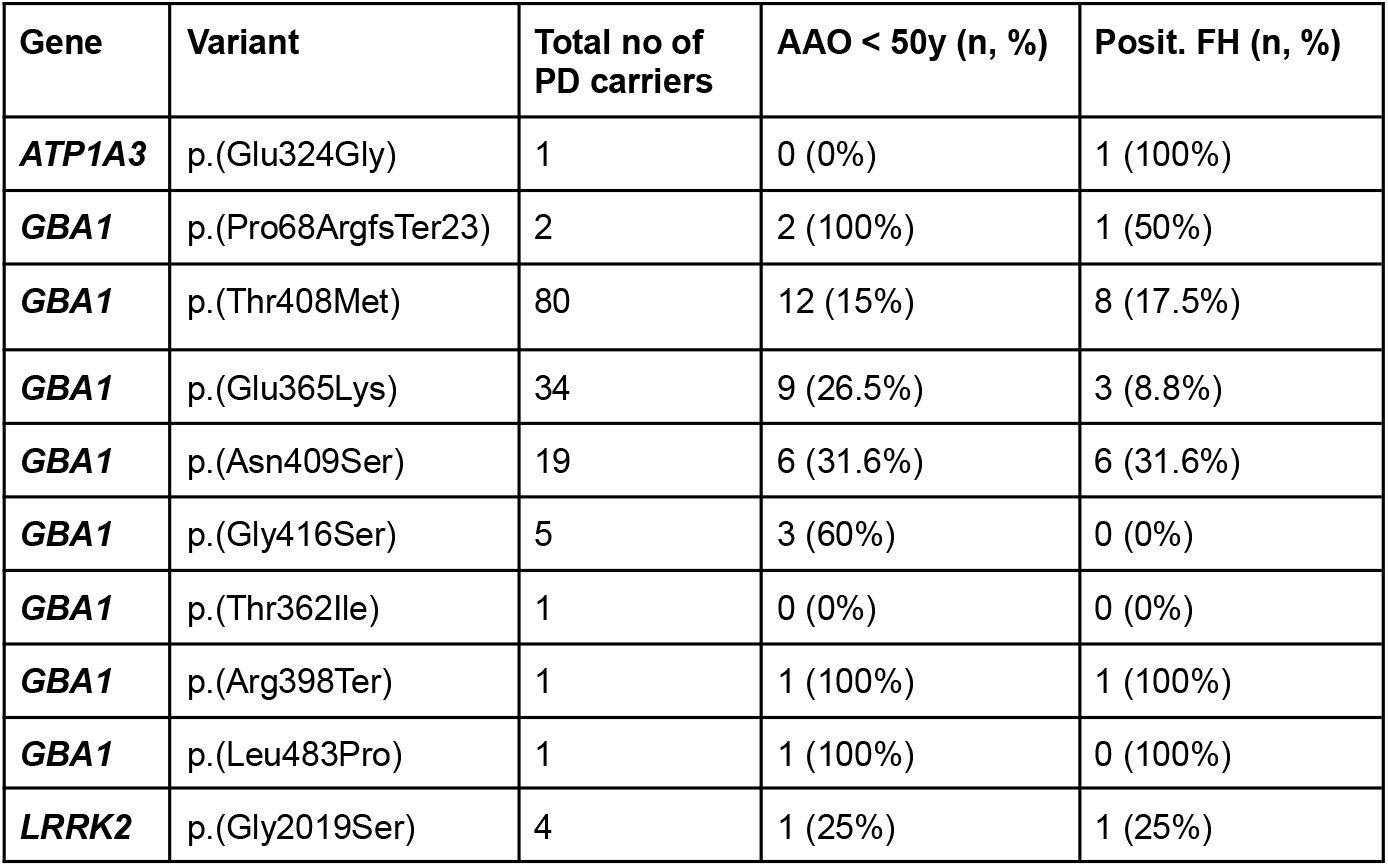
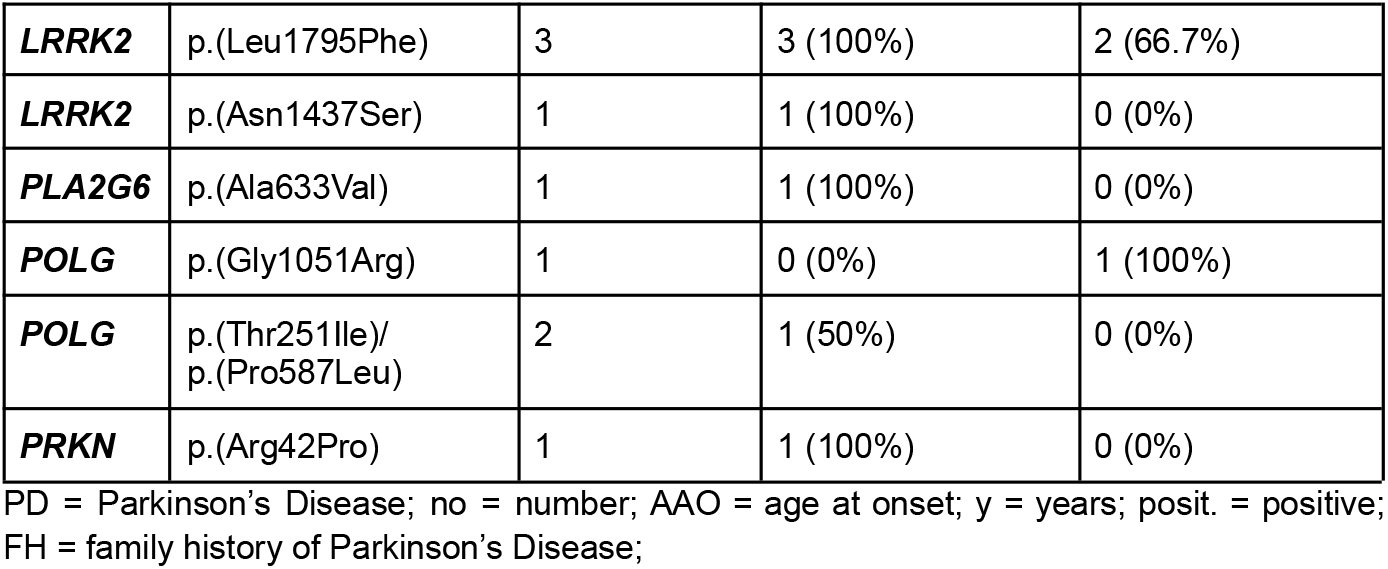
Variant level results: Causative Pathogenic, likely pathogenic or risk factor variants.

## Discussion

The published literature demonstrates a clear evolution of PD genetic testing within the V4 countries over the past two decades. However, initial studies were largely based on PCR and MLPA or Sanger sequencing methods (35/60; 58.3%), focusing on one or a few established PD genes, such as *PRKN, LRRK2* or *GBA1*. While these approaches successfully identified several pathogenic variants, the overall diagnostic yield remained relatively low, usually below 5%, reflecting the limitations of these approaches. With the adoption of the NGS, testing strategies shifted from single-gene analyses to multigene PD panels and WES, which substantially increased the proportion of genetically diagnosed patients to 10-15%, even reaching to 30% in pre-selected high risk cohort of EOPD (Supp. Table 5). The majority of those genetic studies looked at many target genes and reported several rare variants, however many of those currently do not satisfy the stringency of ACMG criteria for reporting in clinical settings, and more evidence is needed to classify those variants as disease-causing.

Another important finding is the unequal geographic distribution of published evidence. Poland accounted for almost half of all identified country-specific studies (19/60; 31.7%), whereas considerably fewer investigations originated from Hungary (8/60; 13.3%), the Czech Republic (8/60; 13.3%) and Slovakia (3/60; 5%). Multicentre collaborations have also played an important role (11/60; 18.3%) (Supp. Table 5).

Therefore, in this largest Central European PD cohort, we aimed to evaluate the diagnostic yield and prevalence of monogenic PD carriers within the region of Central European V4 countries using genotyping and WES data for their advantage to cover large numbers of targets. We focused on carefully curated evidence-based PD-associated virtual gene panel, as evaluated by published literature and Genomics England (Martin et al. 2019) (Supp. Table 4). Overall, we demonstrated a diagnostic yield of reportable genetic variants in 11.4% of enrolled participants. This result is consistent with the previous genetic studies of PD, where 5-10% are estimated to be contributed to monogenic causes. As expected, the *GBA1* risk variants were the most common findings in our cohort, being responsible for nearly 90% (141/157) of all reported variants. However, it is mostly driven by the two most common *GBA1:*p.(Thr408Met) (80/141; 56.7%; monoallelic in 77 cases and bi-allelic in 3 cases) and *GBA1:*p.(Glu365Lys) (34/141 as monoallelic; 24.1%) risk variants, which are currently classified as mild common risk variants (Parlar et al. 2023), and are not generally recommended to disclose for genetic counseling due to their low penetrance (Cook et al., 2024). Although the latest evidence has shown that heterozygous carriers had markedly higher odds of PD than non-carriers across populations, incoming *GBA1*-PD specific clinical trials are opening again the question about disclosing this variant to the patients (Jurecka et al., 2026). Apart from these two variants, we identified six additional *GBA1* pathogenic variants in 29 patients (29/141; 20.6%). Interestingly, the p.(Asn409Ser) variant was the most frequent among the strong *GBA1* risk variants (19/141; 13.5% of all *GBA1* positive cases) identified in our cohort, followed by p.(Gly416Ser) variant, which was present in 3.5% (5/141) of all *GBA1*-PD cases. The p.(Thr362Ile), p.(Arg398Ter) and p.(Leu483Pro) were rare in this cohort; being responsible for only 0.7% (1/141) *GBA1*-PD per each variant respectively. Both the p.(Thr362Ile) and (p.Arg398Ter) variants are considered severe and quite rare (Parlar et al. 2023). The p.(Thr362Ile) was described in European PD cohort in 0.09% of all cases (Lesage et al.2011), while the p.(Arg398Ter) is more associated with South Asian population (Parlar et al.2023), being reported in one EOPD from China (Zhao et al. 2020). The heterozygous deletion c.203del p.(Pro68ArgfsTer23) was detected in two EOPD patients. The deletion, if homozygous, is annotated as pathogenic in Gaucher disease, however, its role in PD is currently unknown (Parlar et al. 2023), and therefore was not disclosed to patients as disease-causing. Carriers of *GBA1* variants have an increased risk of developing PD with earlier onset and possibly a faster motor and non-motor disease progression, including worse cognitive decline, REM sleep disorder and psychiatric symptoms as dominating signs as well (Iwaki et al. 2019). However, for counselling purposes, it is important to acknowledge the existence of large variations in genotype-phenotype correlations and therefore the low predictability for an individual patient (den Heijer et al. 2021).

The second most common monogenic form of PD in this cohort was the LRRK2-associated PD. *LRRK2* mutations accounted for 5.1% (8/157) of all variants reported. Interestingly, the extensively investigated p.(Gly2019Ser) mutation was found only in three patients of Central EU ancestry (as 1 identified carrier was subsequently found to be of Libyan origin), the same number as the newly identified p.(Leu1795Phe) (Ostrozovicova et al., 2025; Lange et al., 2025). Although this comparison should be interpreted with caution, as p.(Leu1795Phe) was identified by WES (n=219), whereas p.(Gly2019Ser) was screened by targeted genotyping in a substantially larger cohort (n=1373), therefore these findings suggest that p.(Leu1795Phe) may be even more prevalent in Central European populations than is currently recognized (Ostrozovicova et al., 2025; Lange et al. 2025) and more large-scale studies are needed to identify its full role in PD within this region. The p.(Asn1437Ser) variant, that seems to occur mostly in German population (Brockman et al., 2011), was identified in one Polish PD patient. In comparison, previous studies in Central Europe showed that *LRRK2* pathogenic variants are quite rare within PD cohorts, with the p.Gly2019Ser variant’s combined prevalence being reported as only 0.33% (MAF = 0.00164) The studied p.(Arg1441His), p.(Asn1437His), p.(Tyr1699Cys), p.(Ile2020Thr), p.(Met1869Val), p.(Pro755Leu), p.(Ile1122Val), p.(Leu1114=), p.(Gly2385Arg) and p.(Arg1428Pro) were not identified at all (Skorvanek et al. 2021).

Regarding the rare Mendelian PD-associated genes, we identified three PD carriers of *POLG* homozygous or double heterozygous mutation, one carrier of *ATP1A3* heterozygous mutation, one carrier of *PLA2G6* homozygous mutation and one carrier of homozygous *PRKN* mutation (6/157; 3.8% of all reported variants). The phenotypic spectrum of *POLG* mutations is broad, however, several patients have been already described to also exhibit signs of levodopa-responsive early-onset parkinsonism as their main clinical presentation in combination with peripheral neuropathy, or progressive external ophthalmology and optic atrophy (Davidzon et al., 2006; Ma et al., 2020; Borsche et al. 2021). In our cohort, we identified one homozygous p.(Gly1051Arg) late-onset, familial PD patient from Slovakia. This variant was linked to POLG-related disease in compound heterozygous state with the p.Arg309His mutation, described in two Italian siblings with neuropathy, ataxia and achalasia, in combination with epilepsy and developmental delay in one sibling, though without signs of parkinsonism (Da Pozzo et al. 2017). Unfortunately, the patient was deceased at the time of this finding, and therefore not available for additional testing. Additionally, we identified two late-onset PD patients (one from Poland and one from Slovakia) as double heterozygous for the *POLG* p.Pro587Leu and p.Thr251Ile variants. The same double heterozygous carrier was also described in literature in one Hungarian PD patient (Illés et al. 2020). Generally, the *POLG*:p.Pro587Leu/p.Thr251Ile are associated to each other, as they exhibit extreme linkage disequilibrium (LD), meaning they are almost always inherited together on the exact same chromosome (in cis) to form a complex allele, and recent studies shown no difference in their frequency in PD cases vs controls, doubting their role in PD (Tay et al., 2026). Therefore, this variant was not disclosed to patients as disease-causing at this stage.

Only one carrier of the pathogenic *ATP1A3* mutation, the p.(Glu324Gly) was detected in heterozygous state in one Slovak familial LOPD patient. Pathogenic variants in this gene have been implicated in several phenotypes in the last decades, usually with an acute onset and paroxysmal episodes triggered by fever or other factors. The first three syndromes described were rapid-onset dystonia parkinsonism, alternating hemiplegia of childhood and cerebellar ataxia, pes cavus, optic atrophy, and sensorineural hearing loss (CAPOS syndrome) (Brashear et al. 2018). However, since their original description, an expanding number of cases presenting with atypical and overlapping features have been reported. Because of this, ATP1A3-disorders are now beginning to be viewed as a phenotypic continuum with broad and heterogeneous clinical symptoms including parkinsonism (Salles et al. 2021). No signs of dystonia were reported in this patient, though available clinical data were limited and therefore closer re-evaluation of the clinical presentation is needed to carefully assess the genotype-phenotype correlation.

The *PLA2G6* biallelic mutations cause early-onset parkinsonism with dystonia, as well as neurodegeneration with brain iron accumulation (Deng et al. 2023). We identified one homozygous EOPD carrier of *PLA2G6:*p.(Ala633Val) variant, which was also present in one published patient, who was compound heterozygous with the p.(Ala341Thr) (Magrinelli et al. 2022), and in one Romanian patient, similarly as a compound heterozygous, though in this case with *PLA2G6* deletion (Ozes et al. 2017). The phenotype of our patient was predominantly as levodopa-responsive early-onset parkinsonism with myoclonus, neuropsychiatric comorbidities and autonomic dysfunction (urine retention and GIT dysfunction). The DaT scan showed reduction in dopaminergic neurons typical for neurodegenerative parkinsonism, while brain MRI revealed classic brain iron accumulation in ncl. caudati, proposing this mutation as disease-causing.

The biallelic *PRKN* mutation was found in only one EOPD patient, carrying the p.(Arg42Pro) known pathogenic mutation. The *PRKN* mutations are considered as the most common recessive cause of PD, however, their identification at a population scale remains challenging, as roughly half are copy number variants (CNVs) in homozygous, or compound heterozygous state (Zhu et al., 2022). A limitation of this study is the lack of systematic CNVs screening, which may have resulted in missed second pathogenic alleles, especially in heterozygous carriers, and further testing is required to help identify additional cases.

Patients with EOPD or familial history are generally considered to be at increased risk for genetic predisposition; however, the clinical recommendations are lacking (Pal et al., 2023). In our cohort, the overall diagnostic yield of genetic testing after completion of both study phases was 11.4% (157/1373 patients), representing an increase from 10.6% (146/1373) after Phase 1 alone. Phase 2 identified an additional 11 pathogenic or likely pathogenic variants among 219 individuals, corresponding to a 5.0% incremental diagnostic yield in the re-tested subgroup. Stratification by clinical characteristics demonstrated a modest enrichment of genetic findings among patients with EOPD (age at onset <50 years), with a diagnostic yield of 12.6% (42/334), compared with 11.1% (115/1039) in those with late-onset disease (≥50 years), representing a relative increase of approximately 14%. Similarly, patients with a positive family history exhibited a diagnostic yield of 12.3% (24/195), which was approximately 11% higher than that observed in the overall cohort. Despite these trends, the differences in diagnostic yield between EOPD, familial cases, and the unselected cohort were relatively modest (approximately 1–1.5 percentage points). Similar observations were already shown by a recent study of 25 063 individuals with PD, where among patients with onset after age 60 years, roughly 1 in 10 to 1 in 12 tested individuals carried a pathogenic or likely pathogenic variant (Balck et al., 2026).These findings suggest that while early age at onset and a positive family history remain useful indicators of an increased likelihood of identifying a genetic cause, clinically actionable variants are also detected in patients without these traditional risk factors. Many patients with *LRRK2*- or *GBA1*-associated PD first present with motor symptoms in their 60s or 70s, and therefore would not be offered testing, even though they carry variants that are the focus of ongoing gene-targeted trials.

## Conclusion

The CEGEMOD study represents the largest assessment of Parkinson’s disease genetics in the V4 countries of Central Europe. We demonstrate that clinically relevant pathogenic or established risk variants are present in over 11% of patients, with *GBA1* and *LRRK2* representing the major contributors, while highlighting the genetic heterogeneity of PD in this previously underrepresented population. These findings improve understanding of the regional genetic architecture of PD and provide evidence supporting broader implementation of genetic testing, particularly in patients with early-onset or familial disease. Together with the publicly available Parkinson Disease Variant Catalogue, this work establishes an important resource for future research, international collaboration, and the development of precision medicine approaches in Parkinson’s disease.

## Data Availability

Anonymised data will be made available upon reasonable request.

## Data Availability

Anonymised data will be made available upon reasonable request.

