## Supplementary for "The Genetic Landscape of Parkinson’s Disease in V4 countries of Central Europe - the CEGEMOD study"

**Supplementary Materials**

**Supp. Table 1: PICOS framework used for Systematic Review**

| **Component** | **Description** |
| --- | --- |
| **Population** | Individuals with PD and HC from Slovakia, the Czech Republic, Poland, and Hungary (Visegrad/V4 countries) |
| **Intervention or Exposure** | Molecular genetic testing of established monogenic Parkinson’s disease genes |
| **Comparison** | Presence and frequency of pathogenic/likely pathogenic/VUS variants or polymorphisms in PD patients compared with healthy controls |
| **Outcome** | Prevalence, distribution, and genetic spectrum of monogenic Parkinson’s disease in V4 populations |
| **Study Type** | Case reports, case series, and observational genetic studies (case–control, cohort, cross-sectional studies) |

PD = Parkinson’s Disease; HC = healthy controls; VUS = variant of unknown significance

**Supp. Table 2: Syntax used for search in Systematic Review**

| **Database** | **Syntax** | **Last searched** | **No**  **of results retrieved** |
| --- | --- | --- | --- |
| MEDLINE/PubMed | parkinson*[Title/Abstract] AND ("gene*"[Title/Abstract] OR "mutation*"[Title/Abstract] OR "variant*"[Title/Abstract] OR "polymorphism*"[Title/Abstract] OR "SNP*"[Title/Abstract] OR "CNV*"[Title/Abstract] OR "copy number variant*"[Title/Abstract] OR "rare variant*"[Title/Abstract] OR "microsatellite*"[Title/Abstract] OR "chromosome*"[Title/Abstract]) AND ("Central Europe"[Title/Abstract] OR "Eastern Europe"[Title/Abstract] OR "Slovak*"[Title/Abstract] OR "Czech"[Title/Abstract] OR "Poland"[Title/Abstract] or "Polish"[Title/Abstract] OR "Hungar*"[Title/Abstract])) | 18/MAY/2026 | 120 |
| Web of Science | (TS=(Parkinson*)) AND (TS=(gene*) OR TS=(mutation*) OR TS=(variant*) OR TS=(polymorphism*) OR TS=(SNP*) OR TS=(CNV*) OR TS=(copy number variant*) OR TS=(rare variant*) OR TS=(microsatellite*) OR TS=(chromosome*)) AND (TS=(Central Europe*) OR TS=(Eastern Europe*) OR TS=(Slovak*) OR TS=(Czech*) OR TS=(Poland*) OR TS=(Polish*) OR TS=(Hungar*)) | 19/MAY/2026 | 301 |
| Scopus | (TITLE-ABS-KEY  "Parkinson*" AND TITLE-ABS-KEY  ("gene*" OR "mutation*" OR "variant*" OR "polymorphism*" OR "SNP*" OR "CNV*" OR "copy number variant*" OR "rare variant*" OR "microsatellite*" OR "chromosome*") AND (TITLE-ABS-KEY ("Central Europe" OR "Eastern Europe" OR "Slovak*"  OR "Czech" OR "Poland" OR "Polish" OR "Hungar*")) | 18/MAY/2026 | 236 |

No = numbers;

**Supp. Figure 1: PRISMA 2020 flow diagram for the systematic review**

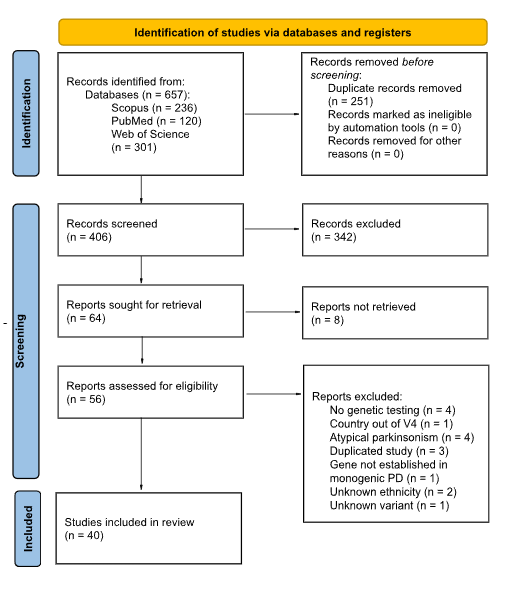

**Supp. Table 3: List of All Eligible Articles Included in the Systematic Review**

| **No** | **Citation** |
| --- | --- |
| 1. | Szlepák T, Kossev AP, Csabán D, Illés A, Udvari S, Balicza P, Borsos B, Takáts A, Klivényi P, Molnár MJ. GBA-associated Parkinson's disease in Hungary: clinical features and genetic insights. Neurol Sci. 2024 Jun;45(6):2671-2679. doi: 10.1007/s10072-023-07213-w. Epub 2023 Dec 28. PMID: 38153678; PMCID: PMC11082009. |
| 2. | Turski P, Chaberska I, Szukało P, Pyska P, Milanowski Ł, Szlufik S, Figura M, Hoffman-Zacharska D, Siuda J, Koziorowski D. Review of the epidemiology and variability of LRRK2 non-p.Gly2019Ser pathogenic mutations in Parkinson's disease. Front Neurosci. 2022 Sep 20;16:971270. doi: 10.3389/fnins.2022.971270. PMID: 36203807; PMCID: PMC9530194. |
| 3. | Kolarikova K, Vodicka R, Vrtel R, Stellmachova J, Prochazka M, Mensikova K, Bartonikova T, Furst T, Kanovsky P, Geryk J. High-Throughput Sequencing Haplotype Analysis Indicates in LRRK2 Gene a Potential Risk Factor for Endemic Parkinsonism in Southeastern Moravia, Czech Republic. Life (Basel). 2022 Jan 14;12(1):121. doi: 10.3390/life12010121. PMID: 35054514; PMCID: PMC8780375. |
| 4. | Dulski J, Cerquera-Cleves C, Milanowski L, Kidd A, Sitek EJ, Strongosky A, Vanegas Monroy AM, Dickson DW, Ross OA, Pentela-Nowicka J, Sławek J, Wszolek ZK. Clinical, pathological and genetic characteristics of Perry disease-new cases and literature review. Eur J Neurol. 2021 Dec;28(12):4010-4021. doi: 10.1111/ene.15048. Epub 2021 Aug 26. PMID: 34342072; PMCID: PMC9295182. |
| 5. | Toth-Bencsik R, Balicza P, Varga ET, Lengyel A, Rudas G, Gal A, Molnar MJ. New Insights of Phospholipase A2 Associated Neurodegeneration Phenotype Based on the Long-Term Follow-Up of a Large Hungarian Family. Front Genet. 2021 Jun 8;12:628904. doi: 10.3389/fgene.2021.628904. PMID: 34168672; PMCID: PMC8217829. |
| 6. | Milanowski ŁM, Lindemann JA, Hoffman-Zacharska D, Soto-Beasley AI, Barcikowska M, Boczarska-Jedynak M, Deutschlander A, Kłodowska G, Dulski J, Fedoryshyn L, Friedman A, Jamrozik Z, Janik P, Karpinsky K, Koziorowski D, Krygowska-Wajs A, Jasińska-Myga B, Opala G, Potulska-Chromik A, Pulyk A, Rektorova I, Sanotsky Y, Siuda J, Sławek J, Śmiłowska K, Szczechowski L, Rudzińska-Bar M, Walton RL, Ross OA, Wszolek ZK. Frequency of mutations in PRKN, PINK1, and DJ1 in Patients With Early-Onset Parkinson Disease from neighboring countries in Central Europe. Parkinsonism Relat Disord. 2021 May;86:48-51. doi: 10.1016/j.parkreldis.2021.03.026. Epub 2021 Apr 2. PMID: 33845304; PMCID: PMC8192481. |
| 7. | Skorvanek M, Rizig M, Athanasiou-Fragkouli A, Necpal J, Straka I, Tamas G, Kurca E, Mosejova A, Han V, Lorincova T, Ostrozovicova M, Liesenerova S, Levicka P, Fajcikova L, Minar M, Valkovic P, Mákos O, Kelemen A, Grofik M, Cibulka M, Jama F, Houlden H; members of the CEGEMOD study group. LRRK2 mutations in Parkinson's disease patients from Central Europe: A case control study. Parkinsonism Relat Disord. 2021 Feb;83:110-112. doi: 10.1016/j.parkreldis.2020.12.021. Epub 2021 Jan 11. PMID: 33561776. |
| 8. | Milanowski Ł, Sitek EJ, Dulski J, Cerquera-Cleves C, Gomez JD, Brockhuis B, Schinwelski M, Kluj-Kozłowska K, Ross OA, Sławek J, Wszolek ZK. Cognitive and behavioral profile of Perry syndrome in two families. Parkinsonism Relat Disord. 2020 Aug;77:114-120. doi: 10.1016/j.parkreldis.2020.05.019. Epub 2020 Jun 22. PMID: 32717578. |
| 9. | Illés A, Balicza P, Gál A, Pentelényi K, Csabán D, Gézsi A, Molnár V, Molnár MJ. Az örökletes Parkinson-kór mint a POLG-gén károsodásának új klinikai megjelenési formája [Hereditary Parkinson’s disease as a new clinical manifestation of the damaged POLG gene]. Orv Hetil. 2020 May 1;161(20):821-828. Hungarian. doi: 10.1556/650.2020.31724. PMID: 32364361. |
| 10. | Milanowski Ł, Hoffman-Zacharska D, Geremek M, Friedman A, Figura M, Koziorowski D. The matter of significance - Has the p.(Glu121Lys) variant of TOR1A gene a pathogenic role in dystonia or Parkinson disease? J Clin Neurosci. 2020 Feb;72:501-503. doi: 10.1016/j.jocn.2019.12.018. Epub 2019 Dec 28. PMID: 31892495. |
| 11. | Vodicka, R., Kolarikova, K., Vrtel, R., Mensikova, K., Kanovsky, P., Prochazka, M. (2020). Evaluating Basic Next-Generation Sequencing Parameters in Relation to True/False Positivity Findings of Rare Variants in an Isolated Population from the Czech Republic South-Eastern Moravia Region with a High Incidence of Parkinsonism. In: Rojas, I., Valenzuela, O., Rojas, F., Herrera, L., Ortuño, F. (eds) Bioinformatics and Biomedical Engineering. IWBBIO 2020. Lecture Notes in Computer Science(), vol 12108. Springer, Cham. https://doi.org/10.1007/978-3-030-45385-5_50 |
| 12. | Boros FA, Török R, Vágvölgyi-Sümegi E, Pesei ZG, Klivényi P, Vécsei L. Assessment of risk factor variants of LRRK2, MAPT, SNCA and TCEANC2 genes in Hungarian sporadic Parkinson's disease patients. Neurosci Lett. 2019 Jul 27;706:140-145. doi: 10.1016/j.neulet.2019.05.014. Epub 2019 May 11. PMID: 31085292. |
| 13. | Illés A, Csabán D, Grosz Z, Balicza P, Gézsi A, Molnár V, Bencsik R, Gál A, Klivényi P, Molnar MJ. The Role of Genetic Testing in the Clinical Practice and Research of Early-Onset Parkinsonian Disorders in a Hungarian Cohort: Increasing Challenge in Genetic Counselling, Improving Chances in Stratification for Clinical Trials. Front Genet. 2019 Oct 31;10:1061. doi: 10.3389/fgene.2019.01061. PMID: 31737044; PMCID: PMC6837163. |
| 14. | Bartoníková T, Menšíková K, Kolaříková K, Vodička R, Vrtěl R, Otruba P, Kaiserová M, Vaštík M, Mikulicová L, Ovečka J, Šáchová L, Dvorský F, Krša J, Jugas P, Godava M, Bareš M, Janout V, Hluštík P, Procházka M, Kaňovský P. New endemic familial parkinsonism in south Moravia, Czech Republic and its genetical background. Medicine (Baltimore). 2018 Sep;97(38):e12313. doi: 10.1097/MD.0000000000012313. PMID: 30235682; PMCID: PMC6160209. |
| 15. | Konno T, Ross OA, Teive HAG, Sławek J, Dickson DW, Wszolek ZK. DCTN1-related neurodegeneration: Perry syndrome and beyond. Parkinsonism Relat Disord. 2017 Aug;41:14-24. doi: 10.1016/j.parkreldis.2017.06.004. Epub 2017 Jun 12. PMID: 28625595; PMCID: PMC5546300. |
| 16. | Bartonikova T, Mensikova K, Mikulicova L, Vodicka R, Vrtel R, Godava M, Vastik M, Kaiserova M, Otruba P, Dolinova I, Nevrly M, Kanovsky P. Familial atypical parkinsonism with rare variant in VPS35 and FBXO7 genes: A case report. Medicine (Baltimore). 2016 Nov;95(46):e5398. doi: 10.1097/MD.0000000000005398. PMID: 27861377; PMCID: PMC5120934. |
| 17. | Török R, Zádori D, Török N, Csility É, Vécsei L, Klivényi P. An assessment of the frequency of mutations in the GBA and VPS35 genes in Hungarian patients with sporadic Parkinson's disease. Neurosci Lett. 2016 Jan 1;610:135-8. doi: 10.1016/j.neulet.2015.11.001. Epub 2015 Nov 4. PMID: 26547032. |
| 18. | Ogaki K, Koga S, Heckman MG, Fiesel FC, Ando M, Labbé C, Lorenzo-Betancor O, Moussaud-Lamodière EL, Soto-Ortolaza AI, Walton RL, Strongosky AJ, Uitti RJ, McCarthy A, Lynch T, Siuda J, Opala G, Rudzinska M, Krygowska-Wajs A, Barcikowska M, Czyzewski K, Puschmann A, Nishioka K, Funayama M, Hattori N, Parisi JE, Petersen RC, Graff-Radford NR, Boeve BF, Springer W, Wszolek ZK, Dickson DW, Ross OA. Mitochondrial targeting sequence variants of the CHCHD2 gene are a risk for Lewy body disorders. Neurology. 2015 Dec 8;85(23):2016-25. doi: 10.1212/WNL.0000000000002170. Epub 2015 Nov 11. PMID: 26561290; PMCID: PMC4676755. |
| 19. | Ambroziak W, Koziorowski D, Duszyc K, Górka-Skoczylas P, Potulska-Chromik A, Sławek J, Hoffman-Zacharska D. Genomic instability in the PARK2 locus is associated with Parkinson's disease. J Appl Genet. 2015 Nov;56(4):451-461. doi: 10.1007/s13353-015-0282-9. Epub 2015 Apr 2. PMID: 25833766; PMCID: PMC4617850. |
| 20. | Oczkowska A, Florczak-Wyspianska J, Permoda-Osip A, Owecki M, Lianeri M, Kozubski W, Dorszewska J. Analysis of PRKN Variants and Clinical Features in Polish Patients with Parkinson's Disease. Curr Genomics. 2015 Aug;16(4):215-23. doi: 10.2174/1389202916666150326002549. PMID: 27006626; PMCID: PMC4765516. |
| 21. | Siuda J, Jasinska-Myga B, Boczarska-Jedynak M, Opala G, Fiesel FC, Moussaud-Lamodière EL, Scarffe LA, Dawson VL, Ross OA, Springer W, Dawson TM, Wszolek ZK. Early-onset Parkinson's disease due to PINK1 p.Q456X mutation--clinical and functional study. Parkinsonism Relat Disord. 2014 Nov;20(11):1274-8. doi: 10.1016/j.parkreldis.2014.08.019. Epub 2014 Sep 2. PMID: 25226871; PMCID: PMC4253017. |
| 22. | Fiala O, Zahorakova D, Pospisilova L, Kucerova J, Matejckova M, Martasek P, Roth J, Ruzicka E. Parkin (PARK 2) mutations are rare in Czech patients with early-onset Parkinson's disease. PLoS One. 2014 Sep 19;9(9):e107585. doi: 10.1371/journal.pone.0107585. PMID: 25238391; PMCID: PMC4169530. |
| 23. | Malec-Litwinowicz M, Rudzińska M, Szubiga M, Michalski M, Tomaszewski T, Szczudlik A. Cognitive impairment in carriers of glucocerebrosidase gene mutation in Parkinson disease patients. Neurol Neurochir Pol. 2014;48(4):258-61. doi: 10.1016/j.pjnns.2014.07.005. Epub 2014 Jul 29. PMID: 25168325. |
| 24. | Hoffman-Zacharska D, Koziorowski D, Ross OA, Milewski M, Poznanski JA, Jurek M, Wszolek ZK, Soto-Ortolaza A, Awek JAS, Janik P, Jamrozik Z, Potulska-Chromik A, Jasinska-Myga B, Opala G, Krygowska-Wajs A, Czyzewski K, Dickson DW, Bal J, Friedman A. Novel A18T and pA29S substitutions in α-synuclein may be associated with sporadic Parkinson's disease. Parkinsonism Relat Disord. 2013 Nov;19(11):1057-1060. doi: 10.1016/j.parkreldis.2013.07.011. Epub 2013 Aug 2. PMID: 23916651; PMCID: PMC4055791. |
| 25. | Koziorowski D, Hoffman-Zacharska D, Sławek J, Jamrozik Z, Janik P, Potulska-Chromik A, Roszmann A, Tataj R, Bal J, Friedman A. Incidence of mutations in the PARK2, PINK1, PARK7 genes in Polish early-onset Parkinson disease patients. Neurol Neurochir Pol. 2013 Jul-Aug;47(4):319-24. doi: 10.5114/ninp.2013.36756. PMID: 23986421. |
| 26. | Bognar C, Baldovic M, Benetin J, Kadasi L, Zatkova A. Analysis of Leucine-rich repeat kinase 2 (LRRK2) and Parkinson protein 2 (parkin, PARK2) genes mutations in Slovak Parkinson disease patients. Gen Physiol Biophys. 2013 Mar;32(1):55-66. doi: 10.4149/gpb2013006. PMID: 23531835. |
| 27. | Balicza P, Bereznai B, Takáts A, Klivényi P, Dibó G, Hidasi E, Balogh I, Molnár MJ. Az LRRK2 gyakori G2019s-mutációjának hiánya 120, korai kezdetu magyar Parkinson-beteg esetében [The absence of the common LRRK2 G2019S mutation in 120 young onset Hungarian Parkinon's disease patients]. Ideggyogy Sz. 2012 Jul 30;65(7-8):239-42. Hungarian. PMID: 23074843. |
| 28. | Gaweda-Walerych K, Safranow K, Jasinska-Myga B, Bialecka M, Klodowska-Duda G, Rudzinska M, Czyzewski K, Cobb SA, Slawek J, Styczynska M, Opala G, Drozdzik M, Nishioka K, Farrer MJ, Ross OA, Wszolek ZK, Barcikowska M, Zekanowski C. PARK2 variability in Polish Parkinson's disease patients--interaction with mitochondrial haplogroups. Parkinsonism Relat Disord. 2012 Jun;18(5):520-4. doi: 10.1016/j.parkreldis.2012.01.021. Epub 2012 Feb 22. PMID: 22361577; PMCID: PMC3358581. |
| 29. | K. Kračunová; M. Kovačovicová; M. Baldovič; P. Valkovič; Ľ. Kádaši; J. Benetin.  The incidence of mutation on the leucine-rich repeat kinase 2 gene in patients with Parkinson's disease in Slovakia \| Výskyt mutácií v gene leucine rich repeat kinase 2 u pacientov s Parkinsonovou chorobou na Slovensku. Cesk Slov Neurol N 2011; 74/107(4): 443-445 |
| 30. | Lesage S, Patin E, Condroyer C, Leutenegger AL, Lohmann E, Giladi N, Bar-Shira A, Belarbi S, Hecham N, Pollak P, Ouvrard-Hernandez AM, Bardien S, Carr J, Benhassine T, Tomiyama H, Pirkevi C, Hamadouche T, Cazeneuve C, Basak AN, Hattori N, Dürr A, Tazir M, Orr-Urtreger A, Quintana-Murci L, Brice A; French Parkinson's Disease Genetics Study Group. Parkinson's disease-related LRRK2 G2019S mutation results from independent mutational events in humans. Hum Mol Genet. 2010 May 15;19(10):1998-2004. doi: 10.1093/hmg/ddq081. Epub 2010 Mar 2. PMID: 20197411. |
| 31. | Koziorowski D, Hoffman-Zacharska D, Sławek J, Szirkowiec W, Janik P, Bal J, Friedman A. Low frequency of the PARK2 gene mutations in Polish patients with the early-onset form of Parkinson disease. Parkinsonism Relat Disord. 2010 Feb;16(2):136-8. doi: 10.1016/j.parkreldis.2009.06.010. Epub 2009 Jul 22. PMID: 19628420. |
| 32. | Fiala O, Pospisilova L, Prochazkova J, Matejckova M, Martasek P, Novakova L, Roth J, Ruzicka E. Parkin mutations and phenotypic features in Czech patients with early-onset Parkinson's disease. Neuro Endocrinol Lett. 2010;31(2):187-92. PMID: 20424582. |
| 33. | Jasinska-Myga B, Wider C, Opala G, Krygowska-Wajs A, Barcikowska M, Czyzewski K, Baker M, Rademakers R, Uitti RJ, Farrer MJ, Ross OA, Wszolek ZK. GRN 3'UTR+78 C>T is not associated with risk for Parkinson's disease. Eur J Neurol. 2009 Aug;16(8):909-11. doi: 10.1111/j.1468-1331.2009.02621.x. Epub 2009 Mar 31. PMID: 19473366. |
| 34. | Białecka M, Hui S, Klodowska-Duda G, Opala G, Tan EK, Droździk M. Analysis of LRRK2 G2019S and I2020T mutations in Parkinson's disease. Neurosci Lett. 2005 Dec 30;390(1):1-3. doi:10.1016/j.neulet.2005.07.012. |
| 35. | Krygowska-Wajs A, Kachergus JM, Hulihan MM, Farrer MJ, Searcy JA, Booij J, Berendse HW, Wolters ECh, Wszolek ZK. Clinical and genetic evaluation of 8 Polish families with levodopa-responsive parkinsonism. J Neural Transm (Vienna). 2005 Nov;112(11):1487-502. doi: 10.1007/s00702-005-0290-8. Epub 2005 Mar 23. PMID: 15785861. |
| 36. | Kachergus J, Mata IF, Hulihan M, Taylor JP, Lincoln S, Aasly J, Gibson JM, Ross OA, Lynch T, Wiley J, Payami H, Nutt J, Maraganore DM, Czyzewski K, Styczynska M, Wszolek ZK, Farrer MJ, Toft M. Identification of a novel LRRK2 mutation linked to autosomal dominant parkinsonism: evidence of a common founder across European populations. Am J Hum Genet. 2005 Apr;76(4):672-80. doi: 10.1086/429256. Epub 2005 Feb 22. PMID: 15726496; PMCID: PMC1199304. |
| 37. | Farrer MJ, Hulihan MM, Kachergus JM, Dachsel JC, Stoessl AJ, Grantier LL, Calne S, Calne DB, Lechevalier B, Chapon F, Tison F, Duhamel A, Destée A, Amouyel P, Wszolek ZK. Two large Polish kindreds with levodopa-responsive Parkinsonism not linked to known Parkinsonian genes and loci. Parkinsonism Relat Disord. 2003 Mar;9(4):193-200. doi:10.1016/S1353-8020(02)00137-5 |
| 38. | Mensikova K, Kanovsky P, Kaiserova M, Vastik M, Hlustik P, Dufek J, Znojil V, Bares M, Vrtel R, Otruba P, Hradilek P, Kralova M, Zapletalova J, Kurcova S, Mikulicova L, Jugas P. Autosomal-dominant parkinsonism with a novel MAPT gene mutation in a specific population of Central Europe (South-Eastern Moravia, Czech Republic). Mov Disord. 2014;29(Suppl 1):S122-S123. |
| 39. | P. Kaňovský, K. Kolaříková, R. Vodička, R. Vrtěl, K. Menšíková, T. Bartoníková, M. Procházka. Rare variants of the LRRK2 gene in the haplotype as one of the potential risk factors for endemic parkinsonism in a small isolated region in the Czech Republic [abstract]. Mov Disord. 2020; 35 (suppl 1). |
| 40. | Kolarikova K, Vodicka R, Vrtel R, Stellmachova J, Prochazka M, Mensikova K, Kanovsky P. Whole Exome Sequencing Study in Isolated South-Eastern Moravia (Czechia) Population Indicates Heterogenous Genetic Background for Parkinsonism Development. Front Neurosci. 2022 Mar 17;16:817713. doi: 10.3389/fnins.2022.817713. PMID: 35368288; PMCID: PMC8968137. |

No = number;

**Supp. Table 4**: List of PD-associated genes screened in the CEGEMOD cohort

| **Source** | **Gene Panel** | **Phase** |
| --- | --- | --- |
| Genomics England Parkinson’s Disease and Complex Parkinsonism panel list v1.120 - green entities | *ATP13A2, ATP1A3, C19orf12, CSF1R, DCTN1, DNAJC6, FBXO7, FTL, GBA1, GCH1, GRN, LRRK2, LYST, MAPT, OPA3, PANK2, PARK7, PDGFB, PINK1, PLA2G6, PRKN, PRKRA, PTRHD1, RAB39B, SLC30A10, SLC39A14 , SLC6A3, SNCA, SPG11, SPR, SYNJ1, TH, TUBB4A, VPS13A, VPS35, WDR45* | *Phase 1; Phase 2* |
| Blauwendrat et al, 2020^1^ | *POLG, DNAJC13, TMEM230, VPS13C, LRP10* | *Phase 1; Phase 2* |
| Gustavsson et al., 2024^2^ | *RAB32* | *Phase 1; Phase 2* |
| Magrinelli et al., 2026^3^ | *PSMF1* | *Phase 1; Phase 2* |

^1^ Blauwendraat C, Nalls MA, Singleton AB. The genetic architecture of Parkinson's disease. Lancet Neurol. 2020 Feb;19(2):170-178. doi: 10.1016/S1474-4422(19)30287-X.

^2^ Gustavsson EK, et al. RAB32 Ser71Arg in autosomal dominant Parkinson's disease: linkage, association, and functional analyses. Lancet Neurol. 2024 Jun;23(6):603-614. doi: 10.1016/S1474-4422(24)00121-2. Epub 2024 Apr 10. PMID: 38614108; PMCID: PMC11096864.

^3^ Magrinelli F, et al. Variants in the proteasome regulator PSMF1 cause a phenotypic spectrum from parkinsonism to perinatal lethality. Nat Commun. 2026 Apr 15. doi: 10.1038/s41467-026-71351-w. Epub ahead of print. PMID: 41986367.

**Supp. Table 5:** Systematic overview of published PD genetic studies from the V4 countries of Central Europe and identified disease-causing or risk factor variants

| **Published study** | **Study group** | **Methods** | **Screening Target** | **No of monogenic PD cases (yield%)** | **Gene: variant identified as pathogenic/risk factor**  **(No, zygosity and ethnicity of PD _carriers)** |
| --- | --- | --- | --- | --- | --- |
| **Czech Republic** | | | | | |
| Fiala et al., 2010 | 45 EOPD | PCR | *PRKN, LRRK2* | 1 (2.22%) | *LRRK2*:p.(Gly2019Ser)  (1 het CZ) |
| Fiala et al., 2014 | 70 EOPD  75 HC | PCR | *PRKN* | 1 (1.43%) | *PRKN*:ex4del  (1 hom CZ) |
| Kanovsky et al. 2014 | 10 PD | NGS | PD gene panel | 0 (0%) | 0 |
| Bartonikova et al. 2016 | case report | NGS gene panel | 16 PD gene panel | 0 (NA) | 0 |
| Bartonikova et al. 2018 | 12 family members (5 PD + 1 PDD + 2 PSP-P + 4 pPD) | PCR | *ADH1C, EIF4G1, FBXO7, GBA + GBAP1, GIGYF2, HTRA2, LRRK2, MAPT, PRKN, DJ-1, PINK1, PLA2G6, SNCA, UCHL1, VPS35* | 0 (0%) | 0 |
| Vodicka et al. 2020 | 30 PD  12 HC | NGS Gene Panel | 16 PD gene panel | 0 (0%) | 0 |
| Kanovsky et al. 2020 | 32 PD  20 HC | NGS Gene Panel | 16 PD gene panel | 0 (0%) | 0 |
| Kolarikova et al. 2022 | 5 family trios | WES | virtual PD gene panel (n=90) | 0 (0%) | 0 |
| **Hungary** | | | | | |
| Várkonyi et al. 2002 | Case report | PCR | *GBA1* | 1 (NA) | *GBA1:*p.(Asn409Ser)  (1 het AJ) |
| Balicza et al., 2012 | 120 PD | PCR | *LRRK2*  (p.Gly2019Ser) | 0 (0%) | 0 |
| Török et al. 2016 | 124 PD (67 as EOPD) 122 HC | PCR-RFLP | *GBA1* (p.Asn409Ser; p.Leu483Pro; p.Arg159Trp); *VPS35* (p.Asp620Asn) | 3 (2.42%) | *GBA1*: p.(Leu483Pro) (3 het HU) |
| Illés et al. 2019 | 142 EOPD (33 as familial) 55 HC | Sanger sequencing (n=142); 127 PD gene panel (n = 40); WES (n=14) | Combination of gene panel (127 PD genes) and WES | 19 (13.38%) | *PRKN:*ex7dup (4 hom HU);  *SNCA*:dup (1 het HU);  *GBA1*:p.(Leu483Pro) (1 het HU);  *GBA1*: p.(His294Gln) (1 het HU):  *GBA1:* p.(Glu365Lys) (2 het HU);  *GBA1:* p.(Thr408Met) (5 het HU);  *GBA1*: p.(Asn409Ser) (2 het HU) |
| Boros et al. 2019 | 124 PD (68 as EOPD)  128 HC | TaqMan and RFLP | *LRRK2, MAPT, SNCA,TCEANC2* | 0 (0%) | 0 |
| Illés et al. 2020 | 67 PD | NGS | *POLG* | 1 (1.49%) | *POLG:*p.(Thr251Ile/p.Pro587Leu)  (1 comp.het HU) |
| Toth-Bencsik et al. 2021 | Case report | Sanger | *PLA2G6*  *(coding region)* | 3 (NA) | *PLA2G6:*p.(Pro622Ser)/p.(Arg600Trp)  (3 comp.het HU) |
| Szlepák et al. 2023 | 190 PD (46 familial) | Sanger (n=36), NGS gene panel (n=138) or WES (n=14) | Combination of targeted screening (*GBA1, PRKN, DJ-1, PINK1, LRRK2, SNCA) and WES* | 32 (16.84%) | *GBA1:* p.(Thr408Met) (16 het HU);  *GBA1:*p.(Glu365Lys) (6 het HU);  *GBA1:* p.(Leu483Pro) (4 het HU);  *GBA1*: p.(Asn409Ser) (3 het HU);  *GBA1*: p.(His294Gln) (2 het HU);  *GBA1*: RecNcil (1 het HU) |
| **Poland** | | | | | |
| Krygowska-Wajs et al. 2003 | Case series | PCR | *SNCA, PRKN, TAU, SCA2, SCA3* | 0 (NA) | 0 |
| Krygowska-Wajs A et al., 2005 | Case series | PCR, MLPA, genotyping | *SCA2, SCA3, SNCA, PRKN, PINK1, DJ-1, MAPT, LRRK2* | 0 (NA) | 0 |
| Kachergus et al. 2005 | Case report | TaqMan | *LRRK2* | 1 (NA) | *LRRK2*:p.(Gly2019Ser)  (1 het PL) |
| Bialecka et al. 2005 | 174 PD (21 as familial; 70 as EOPD) 190 HC | PCR | *LRRK2*  (exon 41) | 0 (0%) | 0 |
| Koziorowski et al. 2009 | 88 EOPD | PCR,  MLPA | *PRKN*  *DJ-1* | 3 (3.4%) | NA |
| Koziorowski et al. 2010 | 79 EOPD (16 as familial) 204 HC | MLPA | *PRKN*  (exons) | 3 (3.8%) | *PRKN.*p.(Q44fsX48)  (1 hom PL);  *PRKN:*p.(Q44fsX48)/ex 4_7del (1 comp.het PL);  *PRKN:*p.(Q44fsX48)/ex 4_7del  (1 comp.het PL);  *PRKN*:ex2dup (1 hom PL) |
| Vilarino Güell et al. 2011 | 362 PD 346 HC | WES | *VPS35* | 0 (0%) | 0 |
| Gaweda-Walerych K et al. 2012 | 104 EOPD  326 PD  315 HC | PCR,  MLPA | *PRKN (exons;*  *104 EOPD)*  *PRKN (S167N, V380L, D394N;*  *326 PD and 315 HC)* | 0 (0%) | 0 |
| Koziorowski et al. 2013 | 150 EOPD 230 HC | PCR, MLPA | *PRKN, PINK1, DJ-1* | 6 (4%) | *PRKN:* Ex3del/Ex4_7del  (1 hom PL);  *PRKN:*Ex2_5dup/p.(Lys211Asn)  (1 comp.het PL);  *PRKN:*Ex3_4del/p.(Gln34ArgfsX5)  (1 comp.het PL);  *PRKN:*Ex4_7del/p.(Gln34ArgfsX5)  (1 comp.het PL);  *PRKN*:Ex2_5dup/p.(Lys211Asn)  (1 comp.het PL);  *PINK1:* p.(Ile3658Asn) (1 hom PL) |
| Hoffman Zacharska et al. 2013 | 629 PD (169 EOPD) 630 HC | PCR | *SNCA* | 0 (0%) | 0 |
| Siuda et al. 2014 | Case report | PCR | *PINK1* | 2 (NA) | *PINK1:* p.(Gln456Ter)  (2 hom PL) |
| Malec-Litwinowics et al. 2014 | 138 PD | PCR | *GBA1*  (ex 8 and 9) | 7 (5%) | *GBA1:*p.(Asn409Ser) (5 het PL)  *GBA1:*p.(Thr408Met) (11 het PL) |
| Lorenzo Betancor et al. 2015 | 101 PD | PCR | *DNAJC13* | 0 (0%) | 0 |
| Oczkowska et al. 2015 | 90 PD (8 as EOPD) 113 HC | PCR | *PRKN* | 0 (0%) | 0 |
| Ambroziak et al. 2015 | 344 PD (171 as EOPD) | MLPA | *PRKN* | 5 (1.45%) | *PRKN.* Ex3del/p.(Cys446Phe)  (1 comp.het PL);  *PRKN:* Ex4_7del/p.(Gln34Argfs*5)  (1 comp.het PL);  *PRKN:* Ex3del/Ex4_7del  (1 comp.het PL);  *PRKN*: Ex3_4del/p.(Gln34Argfs*5)  (1 comp.het PL);  *PRKN:* Ex2_5dup/p.(Lys211Asn)  (1 comp.het PL) |
| Jamrozik, Lugowska and Kosiorowski 2015 | 270 PD (115 as EOPD) | PCR-RFLP | *GBA1* (p.Leu483Prop.Asn409Ser) | 11 (4.07%) | *GBA1*:p.(Leu483Pro) (4 het PL);  *GBA1*:p.(Asn409Ser) (7 het PL) |
| Konno et al. 2017 | Case report | Clinical genetic testing | *MAPT, GRN, C9orf72, DCTN1* | 1 (NA) | *DCTN1:*p.(Gly71Glu) (1 het PL) |
| Hanna Al Shaikh et al. 2022 | Case report | WES | *PLA2G6* case series | 1 (NA) | *PLA2G6:* p.(Thr661Met)/p.(Ala781Thr) (1 comp.het PL) |
| Turski et al. 2022 | systematic review and case report | WES family analysis | NA | 3 (NA) | *LRRK2:*p.(Asn1437His) (3 het PL) |
| **Slovakia** | | | | | |
| Kracunova et al. 2011 | 126 PD (18 as familial PD; 10 as EOPD | dHPLC | *LRRK2*  (exons 31,35,41,48) *PRKN*  (exons) | 0 (0%) | 0 |
| Bognar et al. 2013 | 160 PD | PCR | *SNCA*  (exon 2 and 3) | 0 (0%) | 0 |
| Bognar et al. 2013 | 216 PD | dHPLC | *LRRK2*  (exons 31,35,41,48) *PRKN*  (exons 2,6,7) | 0 (0%) | 0 |
| **Multicentric Studies including PD patients from Central Europe** | | | | | |
| Jasinska-Myga et al. 2009 | PL: 364 PD  PL: 236 HC | PCR | *GRN* (c.*78C>T) | 206 (57%) | NA (Benign variant) |
| Lesage et al. 2010 | additional LRRK2 + families from selected countries | case series | *LRRK2*  *p.(Gly2019Ser)* | 12 (NA) | *LRRK2*:p.(Gly2019Ser)  (10 het PL;  2 het HU) |
| Lorenzo Betancor et al. 2015 | 748 PD (705 PL; 33 CZ; 10 UA) | PCR | *DNAJC13* | 0 (0%) | 0 |
| Ogaki et al. 2015 | 394 PD  (58 as familial)  350 HC | PCR | *CHCHD2* (exons) | 0 (0%) | 0 |
| Schormair et al. 2018 | 80 EOPD | WES | PD-associated virtual gene panel | 22 (27.5%) | *GBA1:*p.(Asn409Ser)  (2 het NA);  *GBA1*: p.(Leu483Pro)  (1 het NA);  *GBA1*: p.(Asp448His)  (1 het NA);  *GBA1*: p.(Trp432Ter)  (1 het NA);  *GBA1:* p.(Glu365Lys)  (10 het NA);  *GBA1:* p.(Thr408Met)  (3 het NA);  *LRRK2*:p.(Gly2019Ser)  (2 het NA);  *PINK1:*p.(Arg492Ter) (1 hom NA);  *PINK1*;ex3-4del (1 hom NA);  *VPS13C*:c.2029+2T>G/c.3215-1G>T  (1 comp.het NA) |
| Milanowski et al. 2020 | case report | genetic testing - not specified | not specified | 1 (NA) | *DCTN1*:p.Gly71Glu (1het PL) |
| Milanowski et al. 2021 | 541 EOPD (11 CZ; 38 DE; 476 PL; 16 UA) | Sanger, MLPA | *PRKN; PINK1; DJ-1* | 19 (3.51%) | *PRKN:*Ex2_4del/Ex3_4del  (1 comp.het CZ);  *PRKN:*p.(Lys211Asn)/p.(Arg275Trp)  (1 comp.het CZ);  *PRKN:*Ex3del/Ex4_7del  (1 comp.het PL);  *PRKN:*Ex4_7del/p.(Gln34ArgfsTer5)  (1 comp.het PL);  *PRKN:*Ex2_5dup/p.(Lys211Asn)  (1 comp.het PL);  *PRKN:*p.(Gln34ArgfsTer5)/p.(Gln34ArgfsTer5) (1 comp.het PL);  *PRKN:*Ex3del/p.(Cys446Phe)  (1 comp.het PL);  *PRKN:*Ex4del/p.(Lys211Asn)  (1 comp.het PL);  *PRKN:*Ex2del/Ex4del  (1 comp.het PL);  *PRKN:*Ex2del/p.(Glu79Ter)  (1 comp.het PL);  *PRKN:*p.(Arg275Trp)/p.(Pro437Leu)  (1 comp.het PL);  *PRKN:*Ex3del/Ex5_9del  (1 comp.het PL);  *PRKN:*Ex3_4del/p.(Gln34ArgfsTer5)  (1 comp.het PL);  *PRKN:*Ex2dup/p.(Gln34ArgfsTer5)  (1 comp.het PL);  *PRKN:*p.(Gln34ArgfsTer5)/p.(Arg275Trp) (1 comp.het UA);  *PINK1*:p.(Ile368Asn) (1 hom PL);  *PINK1*:p.(Ala168Pro) (1hom PL);  *PINK1*:p.(Gln456Ter) (1 hom PL) |
| Skorvanek et al. 2021 | 732 PD (86/732 familial; 83/732 EOPD) 342 HC | KASP | *LRRK2*  (12 SNPs) | 4 (0.55%) | *LRRK2:*p.(Gly2019Ser)  (2 het SVK); |
| Dulski et al. 2021 | systematic review and case report | Direct seq. | *DCTN1* (ex2) | 2 (NA) | *DCTN1:*p.(Gly71Glu) (2 het PL) |
| Ostrozovicova et al. 2025 | 219 EOPD (93/219 familial)  303 HC | WES | *LRRK2*  p.Leu1795Phe | 3 (1.37%) | *LRRK2:*p.(Leu1795Phe)  (2 het SVK; 1 het HU) |

PD = Parkinson’s Disease, EO = Early-Onset; HC = Healthy Control; n = number; PCR = Polymerase Chain Reaction; RFLP = Restriction Fragment Length Polymorphism; PGM = personal genome machine; WES = whole-exome sequencing; NGS = next generation sequencing; MLPA = Multiplex Ligation-dependent Probe Amplification; dHPLC = denaturing High Performance Liquid Chromatography; KASP = Kompetitive Allele Specific PCR; ex = exon; SNPs = Single Nucleotide Polymorphisms; dup = duplication; del = deletion; CZ = Czech; AJ = Ashkenazi Jewish; HU = Hungarian; PL = Polish; SVK = Slovak; UA = Ukrainian; DE = German; NA = not available;
